# Establishment of Reference Values of Markers of Non-Thyroidal Illness Syndrome on a State-of-the-Art Fully Automated ABEI-Based Chemiluminescence System

**DOI:** 10.1101/2024.06.26.24309539

**Authors:** Aamir Ijaz, Muhammad Usman Anjum, Farhan Ahmed, Shamila Najeeb Paracha, Farzana Habib, Muhammad Bilal Shaukat, Sara Najeeb, Najma Hafeez

**Affiliations:** NUST School of Health Sciences Islamabad, Pakistan; Mohi Uddin Islamic Medical College and Hospital Mirpur AJ&K, Pakistan; Milton-Keyenes University Hospital NHS Trust, Oxford University Hospitals UK

**Keywords:** Reverse Triiodothyronine (rT3), Thyroid Stimulating Hormone (TSH), Free Triiodothyronine (fT3) and Free Thyroxine (fT4)

## Abstract

**Background:** Non-Thyroidal Illness Syndrome (NTIS), previously called Euthyroid Sick Syndrome has always been a diagnostic dilemma for Chemical Pathologists and treating physicians. Combination of four test i.e. Reverse Triiodothyronine (rT3), Free Triiodothyronine (fT3), Free Thyroxine (fT4) and Thyroid Stimulating Hormone (TSH) can be helpful to diagnose NTIS in critically ill patients. We need, however, to determine the reference values found in a reference population, free of any thyroidal as well as non-thyroidal illness.

**Methods and Materials:** Initially, 266 adult volunteers were included in the study from Mohi Uddin Islamic Medical College Mirpur AJ&K (MIMC) and Mohi Uddin Teaching Hospital Mirpur AJ&K (MOTH) after taking written informed consent. About 124 volunteers were selected (males 61; females 63) after exclusion of the volunteers with one or more of the exclusion criteria. Blood specimens were collected and analyzed for rT3, fT3, fT4 and TSH using Maglumi X8 (S nibe, China), an ABEI (N-(aminobutil)-N-(ethyl)-isoluminol) based analyzer at Pathology Laboratory MOTH.

**Results:** Reference values of rT3, fT3, fT4 and TSH were found to be 13.3 – 19.8 ng/dL (0.2 – 0.30 nmol/L); 2.65 - 4.15 pg/mL (4.08 - 6.39 pmol/L);10.6 - 16.2 pg/mL (13.64–20.84 pmol/L) and 0.43 - 3.98 μIU/mL (0.43 – 3.98 mIU/L), respectively.

**Conclusion:** Reference values determined in our own population should be used when diagnosing and monitoring NTIS and other thyroid disorders

## Introduction

Non-Thyroidal Illness Syndrome (NTIS), previously called Euthyroid Sick Syndrome is characterized by increased reverse T3 (rT3), and low levels of triiodothyronine (T3), thyroxine (T4) and thyroid-stimulating hormone (TSH) without previous hypothalamic-pituitary dysfunction or preexisting thyroid gland disease^1^. Another hallmark of NTIS is complete normalization of these hormones after recovery of the disease that causes the changes^2^. NTIS is a common event in critically ill patients (44-70%)^1^. Besides the central regulation of thyroid hormones in the serum by the hypothalamic–pituitary–thyroid (HPT) axis, thyroid concentration inside the cells is tightly controlled by three iodothyronine deiodinase enzymes (DIO1, DIO2 and DIO3), which catalyze the removal of iodine atoms at the outer ring (activation pathway-production of T3) or at the inner ring (inactivation pathway – production of rT3) of T4 and T3^3^ (Figure 1). The hormonal changes in NTIS (decreased T3 and increased rT3 and rT3/T4) observed during the acute phase of illness are considered beneficial but it is related to the fasting state of the body^4^. Early enteral feeding does not produce these hormonal changes^5^. They may become deleterious during prolonged critical illness, making the stage and severity of illness a major determinant of NTIS^5^. The list of diseases and conditions in which NTIS can occur is quite long and growing e.g. pneumonia, starvation, anorexia nervosa, sepsis, stress, cardiopulmonary bypass, myocardial infarction, malignancies, burns, organ transplantations, hypothermia, inflammatory bowel disease, cirrhosis, major surgery, renal failure, diabetic ketoacidosis^6,^, fracture of hip joint^7^, congestive cardiac failure^8^ and COVID-19 infection^9^. There may be unusual variations of thyroid function in some conditions e.g. decrease in the reverse T3 and rT3/T4 ratio, and normal T3 and T3/T4 ratio in HIV^10^.

**Figure 1:**
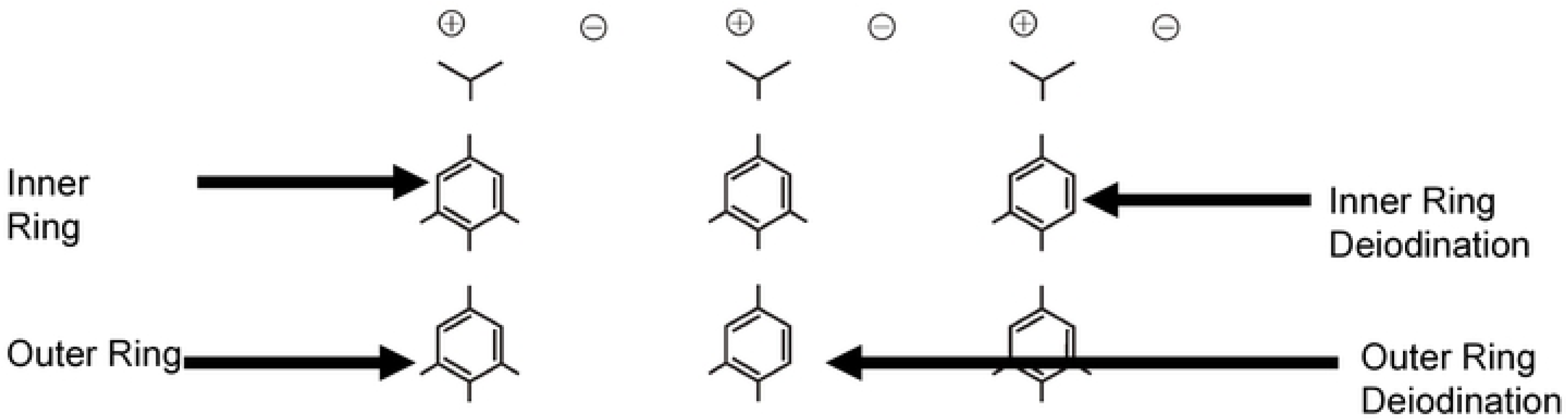
Positions of Iodine Atoms in Thyroid Hormones

We use the term “reference values” because “normal values” and “reference ranges” are obsolete terms now. There are several different connotations of the term “normal” in statistics, epidemiology and clinical practice^11^, while the term “range” implies a single value i.e. difference between maximum and minimum values in a distribution^12^.

International Federation of Clinical Chemistry (IFCC) has also recommended usage of term “reference values”^13^. Reference values can be determined from a set of large subject or patient data (*posteriori)* or by a pre-planned specimen collection from a reference population after careful exclusion of the disease for which the test is used *(priori).* For less commonly used analytes such as the hormones a priori method is preferable^14^. Consideration of biological factors is another important step in reference values studies. The controllable factors are neutralized by standardization of the procedure for preparation of reference individuals and specimen collection, while non-controllable factors such as age and gender become partitioning criteria^15^. Three different statistical methods are used for establishment of reference values i.e. non-parametric methods^16^ e.g. boot-strap method^17^, parametric method^18^ and robust method^19^. All these method have their own inherent advantages and limitations^20^.

### Methods and Material

### Venue of the Project

Mohi Uddin Islamic Medical College, Mirpur AJ&K, Pakistan (MIMC) and Mohi Uddin Teaching Hospital Mirpur AJ&K, Pakistan (MOTH)

### Permission from Institutional Review Board (IRB)

Proper permission was obtained from IRB of MIMC.

### Study Duration

One year starting from 31^st^ August 2022 to 15^th^ August 2023.

### Reference individuals

Two hundred and sixty six volunteers were included in the study, mostly medical students and staff of MIMC and MOTH after taking their informed written consent.

### Exclusion criteria

1. Age < 20 years and > 50 years
2. Mild Diseases
3. Participants on any medications
4. Pregnant participants
5. Chronic Illness
6. Thyroid disorders
7. Menstrual irregularities
8. Infertility

### Sample Size

A total of 266 volunteers were included in the study for specimen collection. This sample size was used keeping in mind the recommended minimum sample size of minimum 120 individuals for establishing reference values without partitioning for gender and gender^20^.

### Pre-analytical Factors

were addressed carefully before blood specimens were collected and analyzed including:.

a. Subject preparation: Volunteers were briefed about the venipuncture to alley their anxiety.
b. Specimen collection: non-fasting condition, sitting relaxed for about 10 minutes prior to the venipuncture, no double prick and use of fine needles to reduce pain.
c. Specimen handling: early transport to the lab for centrifugation and serum separation
d. Specimen Transport: Specimens were immediately transported to the lab
e. Specimen storage: Stored in an appropriate refrigerator until analyzed

### Laboratory Analyses

a. After exclusion of the volunteers meeting the exclusion criteria, blood specimens of 124 volunteers were analyzed for four hormones related to NTIS i.e. rT3, fT3, fT4 and TSH.
b. Chemiluminescence-based autoanalyzer Maglumi-X8 by SNIBE (Shenzhen, China) was used for laboratory based analyses, This, a continuous random access chemiluminescence immunoassay (CLIA) automated system, uses nano-magnetic microbeads separation high throughput (180 tests/h) technology, the luminescence substrate being N-(aminobutil)-N-(ethyl)-isoluminol (ABEI).
c. Snibe (Shenzhen China) provided reagents and consumables for this project.
d. Performance Characteristics of these tests provided by the manufacturers are given in Table 1.
e. All laboratory results were blinded by allotting unique numbers to the volunteers blood specimens. The lab results of outliers, however, were unblinded for repeating and referral to the related physicians for management.

### Quality Control (QC)

a. Internal QC provided in the reagent kits was run with each batch of analysis
b. NHS-EQAS^®^ was participated to ensure accuracy by inter-laboratory comparison.

### Statistical Procedures

1. Non-parametric approach was used for calculation of reference values because of following reasons:

_a._ This is the recommended method by IFCC^13^ and adopted by Clinical and Laboratory Standard Institute (CLSI) of USA^20^
b. The nature of the underlying distribution of the data does not matter.
c. No statistical expertise is required; The values obtained from reference individuals were simply put in rank order by concentration (rank 1 is the lowest, rank 2 is the next lowest, etc.).
d. The central 95% became the reference interval.
e. The 90% confidence limits of the endpoints of the interval was similarly just taken from the data points themselves.
f. One hundred and twenty observations provide enough data to determine both the central 95% of the distribution and the 90% confidence limits on both endpoints. That is, with 120 observations, rank 3 is the 2.5^th^ percentile; rank 118 is the 97.5^th^ percentile; ranks 1 and 7 define the 90% confidence interval of the 2.5^th^ percentile; and ranks 114 and 120 define the 90% confidence interval of the 97.5^th^ percentile^12^.
2. Since no statistical difference between the two partitioned groups e.g. males and females was found, so separate reference intervals for different partitions were not needed and data was not collected for each partition

## Results

One hundred and forty two volunteers were excluded from the study due to falling in one of the exclusion criteria mentioned above (Figure 1). Females (n=41) and Males (n=27) of 20-24 years of age were the most dominant group (Figure 2). Post-analyses exclusion was done for outliers based on Dixon’s Method^12^ (0-3 per analyte) (Table 2). Three results of TSH were too high to be included in the data and declared outliers. One participant had high TSH and low fT4 (Primary Hypothyroidism), while in the other two, only TSH was raised (possibly sub-clinical hypothyroidism). All three were referred to the concerned physicians for proper management. Details of the non-parametric procedure used for the determination of lower and upper reference values along with 90% Confidence Limits of all four hormones are given in Table 2. Table 3 indicates the reference values of 4 hormones in traditional and SI units along with the conversion factors for general utility. In Table 3 we have also compared the reference values determined using Maglumi X8, reference values reported by the manufactures^21–24^ and reference values established using Abbott Architect (US)^25^.

**Figure 2:**
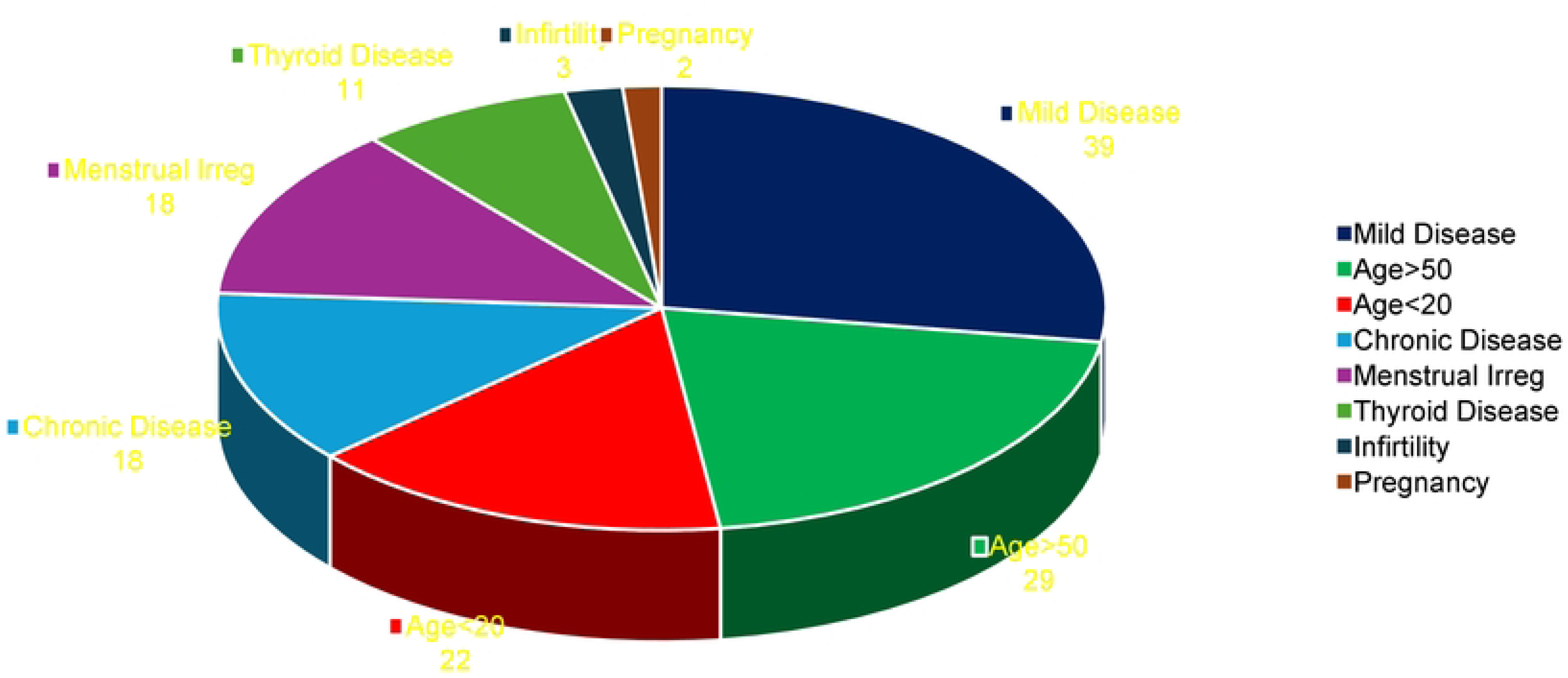
Distribution of Volunteers Excluded from Study (n = 142)

**Figure 3:**
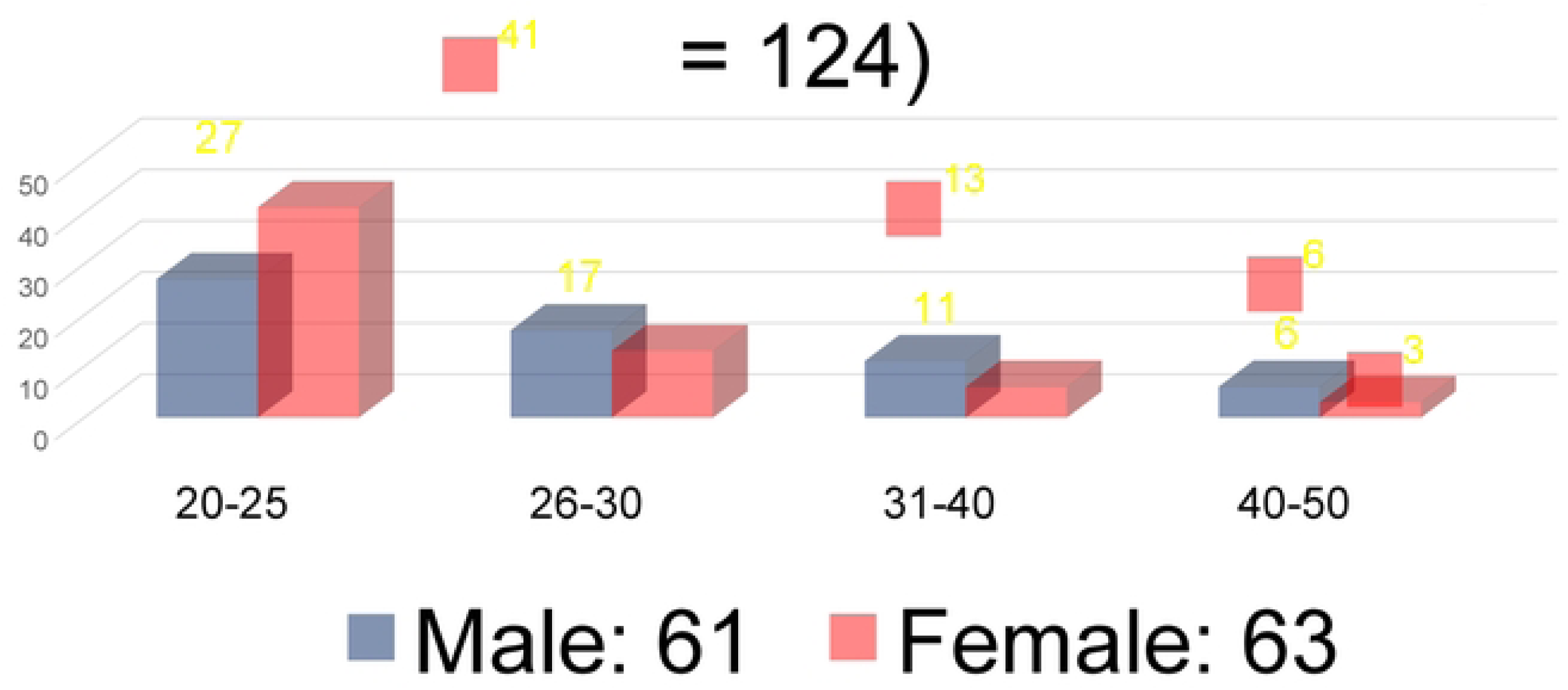
Age and Gender Distribution of Selected Volunteers (n = 124)

**Table 1:** Performance Characteristics of NTIS Related Hormones.

| Name of Hormone | Precision (Intra-assay) CV% | LOB & LOD | Analytical Specificity (Cross reactivity) | AMR | Linearity | Method Comparison |
| --- | --- | --- | --- | --- | --- | --- |
| Reverse T3<sup>21</sup> | 4.88% | 5 & 10 ng/dL | None | 5 – 100 ng/dL | r=0.997 | y=0.985x + 0.007 |
| Free T3<sup>22</sup> | 4.63% | 0.2 & 0.4 pg/mL | T4: <0.01%<br>T3: <0.01 % | 0.2 – 50 pg/mL | r=0.9506 | y= 0.970x + 0.02338 |
| Free T4<sup>23</sup> | 4.24% | 1.0 & 1.5 pg/mL | T4: <0.01%<br>T3: <0.01 % | 1.0 – 120 pg/mL | r=0.997 | y= 1.019x + 0.442 |
| TSH<sup>24</sup> | 2.53% | 0.001 & 0.006 mIU/L | None | 0.001 – 100 mIU/L | r=0.9596 | y=0.953x + 0.327 |

**Table 2:**
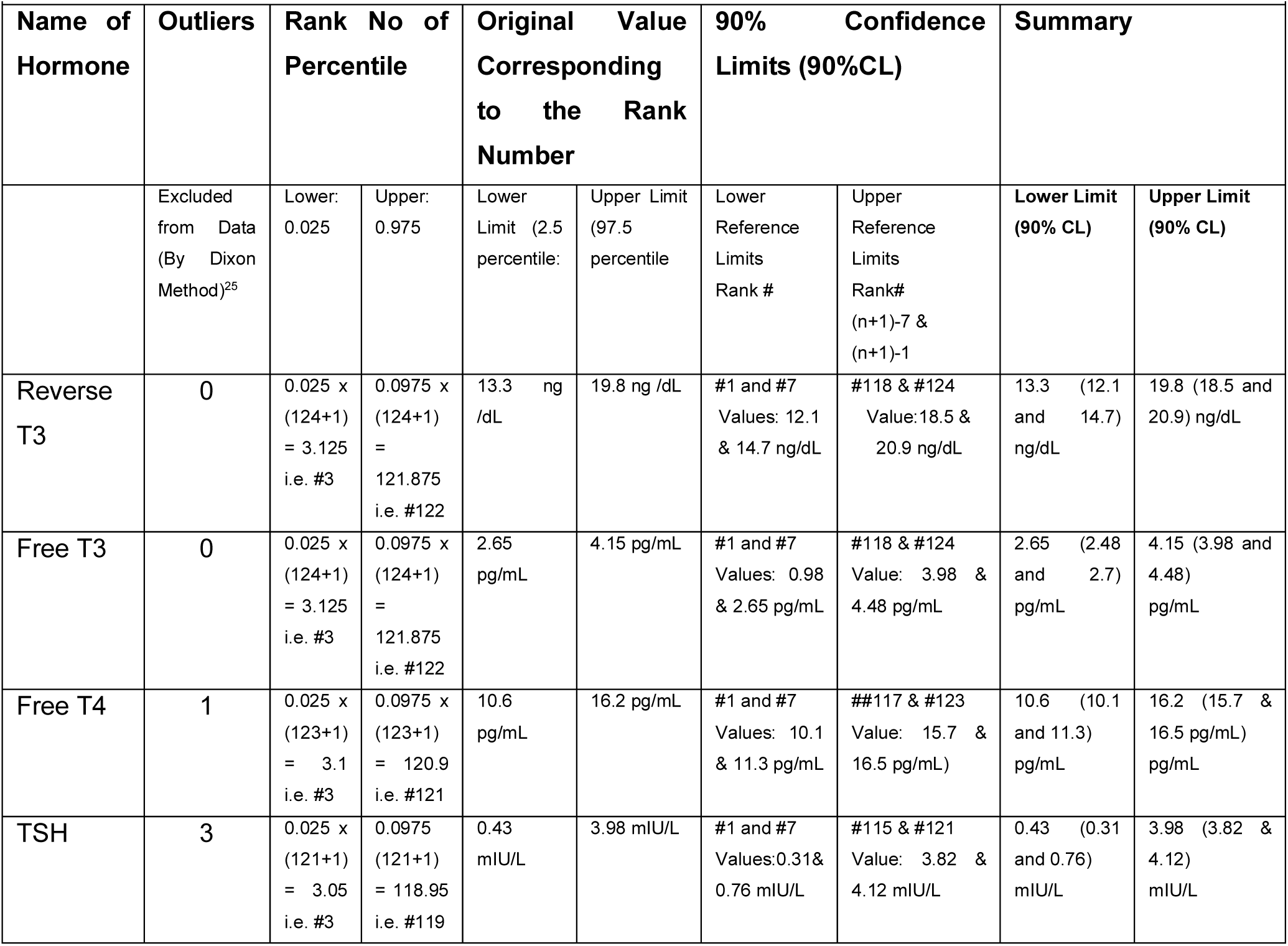
Non-Parametric Determination of Reference Values of NTIS Related Hormones^12^ (Number of Blood Specimen Analyzed = 124)

| Name of Hormone | Outliers | Rank No of Percentile |  | Original Value Corresponding to the Rank Number |  | 90% Confidence Limits (90%CL) |  | Summary |  |
| --- | --- | --- | --- | --- | --- | --- | --- | --- | --- |
|  | Excluded from Data (By Dixon Method)<sup>25</sup> | Lower: 0.025 | Upper: 0.975 | Lower Limit (2.5 percentile) | Upper Limit (97.5 percentile) | Lower Reference Limits Rank # | Upper Reference Limits Rank# (n+1)-7 & (n+1)-1 | Lower Limit (90% CL) | Upper Limit (90% CL) |
| Reverse T3 | 0 | 0.025 x (124+1) = 3.125 i.e. #3 | 0.0975 x (124+1) = 121.875 i.e. #122 | 13.3 ng /dL | 19.8 ng /dL | #1 and #7 Values: 12.1 & 14.7 ng/dL | #118 & #124 Value: 18.5 & 20.9 ng/dL | 13.3 (12.1 and 14.7) ng/dL | 19.8 (18.5 and 20.9) ng/dL |
| Free T3 | 0 | 0.025 x (124+1) = 3.125 i.e. #3 | 0.0975 x (124+1) = 121.875 i.e. #122 | 2.65 pg/mL | 4.15 pg/mL | #1 and #7 Values: 0.98 & 2.65 pg/mL | #118 & #124 Value: 3.98 & 4.48 pg/mL | 2.65 (2.48 and 2.7) pg/mL | 4.15 (3.98 and 4.48) pg/mL |
| Free T4 | 1 | 0.025 x (123+1) = 3.1 i.e. #3 | 0.0975 x (123+1) = 120.9 i.e. #121 | 10.6 pg/mL | 16.2 pg/mL | #1 and #7 Values: 10.1 & 11.3 pg/mL | ##117 & #123 Value: 15.7 & 16.5 pg/mL | 10.6 (10.1 and 11.3) pg/mL | 16.2 (15.7 & 16.5) pg/mL |
| TSH | 3 | 0.025 x (121+1) = 3.05 i.e. #3 | 0.0975 (121+1) = 118.95 i.e. #119 | 0.43 mIU/L | 3.98 mIU/L | #1 and #7 Values: 0.31 & 0.76 mIU/L | #115 & #121 Value: 3.82 & 4.12 mIU/L | 0.43 (0.31 and 0.76) mIU/L | 3.98 (3.82 & 4.12) mIU/L |

**Table 3:**
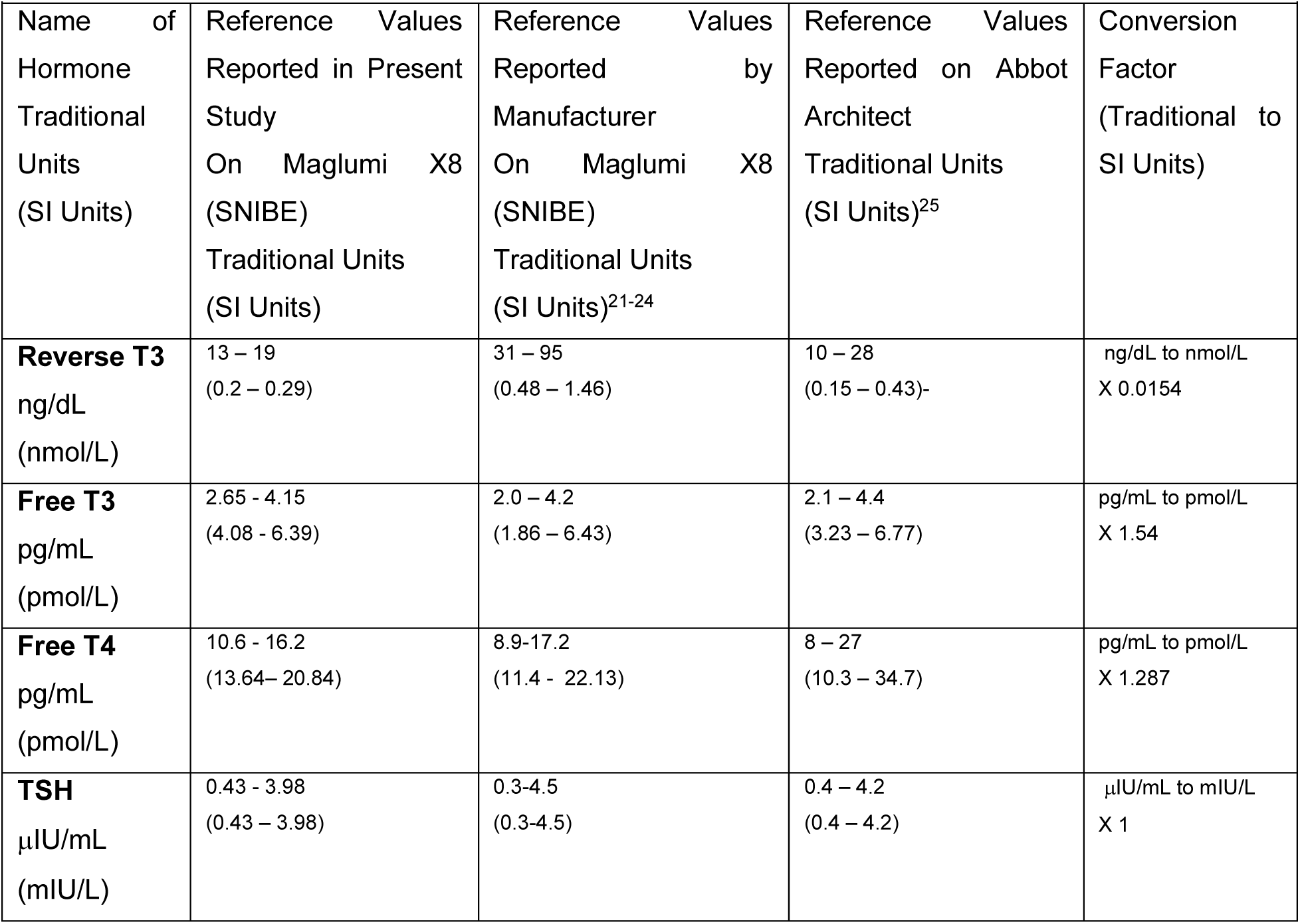
Reference Values NTIS Related Hormones Determined by Different Systems in Traditional and SI Units.

|  |  |  |  |  |
| --- | --- | --- | --- | --- |
| Name of Hormone<br>Traditional Units<br>(SI Units) | Reference Values<br>Reported in Present Study<br>On Maglumi X8 (SNIBE)<br>Traditional Units<br>(SI Units) | Reference Values<br>Reported by Manufacturer<br>On Maglumi X8 (SNIBE)<br>Traditional Units<br>(SI Units)<sup>21-24</sup> | Reference Values<br>Reported on Abbot Architect<br>Traditional Units<br>(SI Units)<sup>25</sup> | Conversion Factor<br>(Traditional to SI Units) |
| **Reverse T3**<br>ng/dL<br>(nmol/L) | 13 – 19<br>(0.2 – 0.29) | 31 – 95<br>(0.48 – 1.46) | 10 – 28<br>(0.15 – 0.43)- | ng/dL to nmol/L<br>X 0.0154 |
| **Free T3**<br>pg/mL<br>(pmol/L) | 2.65 - 4.15<br>(4.08 - 6.39) | 2.0 – 4.2<br>(1.86 – 6.43) | 2.1 – 4.4<br>(3.23 – 6.77) | pg/mL to pmol/L<br>X 1.54 |
| **Free T4**<br>pg/mL<br>(pmol/L) | 10.6 - 16.2<br>(13.64– 20.84) | 8.9-17.2<br>(11.4 - 22.13) | 8 – 27<br>(10.3 – 34.7) | pg/mL to pmol/L<br>X 1.287 |
| **TSH**<br>μIU/mL<br>(mIU/L) | 0.43 - 3.98<br>(0.43 – 3.98) | 0.3-4.5<br>(0.3-4.5) | 0.4 – 4.2<br>(0.4 – 4.2) | μIU/mL to mIU/L<br>X 1 |

## Discussion

This is the first study in Pakistan in which reference values of rT3 has been determined along with other markers of NTIS. The reference values of all four hormones have been compared with reference values determined on another chemiluminescence autoanalyzer i.e. Architect (Abbott)^25^. Values of TSH of two systems are closest followed by fT3, rT3 and fT4 (Table 3). Except for rT3 reference values reported by the manufacturers (Snibe-China)^21–24^ are quite similar to those reported in the present study (Table 3).

rT3 is a unique thyroid hormone which is secreted in an extra-thyroidal site and is used for the exclusion of thyroid diseases^26^. There has been a close contest in the NTIS literature between rT3 and fT3 as diagnostic markers for NTIS, with fT3 mostly winning the contest of diagnostic marker of NTIS^2^. The probable reasons are easy availability and more consistent decrease of fT3 in NTIS, while rT3 and rT3 / fT3 is increased in most of the cases but may be normal or even decreased in some situations^10^. With the advent of better analytical method and equipment (e.g. CLIA and Maglumi by Snibe-China), rT3 is gaining popularity^21^. Moreover, in the presence of central hypothyroidism (pituitary or hypothalamic origin), rT3 will be quite helpful to the physicians. Although role of rT3 in NTIS is questionable, in elderly it has been shown that increased rT3 was the only thyroid hormonal change related to shorter patient survival, with a specificity to predict early death up to 98.5%^27^. Similarity, decreased fT3, fT4 and TSH have also been reported as effective mortality predictors^27^.

It is important to note that NTIS has two faces-acute illness and chronic illness^28^. Divided chronologically, one is good and the other may be bad, depending on the context^5^. A large RCT—the Early versus late Parenteral Nutrition in Critically Ill Adults (EPaNIC) trial— has shown that a macronutrient deficit that occurs in ICU patients during the first week, when only relying on enteral feeding, improves outcomes as compared with preventing that macronutrient deficit with early initiation of parenteral nutrition^29^. Moreover, ‘early fasting’ showed several benefits mediated through hormonal changes of NTIS i.e. increased rT3, decreased fT3, fT4 and TSH e.g. decreased hospital infections, liver abnormalities, muscle weakness, and accelerated recovery^30–32^. It has been implicated that this situation is developed to decrease the metabolic rate of the body and conserve energy^5^. NTIS that occurs in prolonged critically ill patients, who continue to be dependent on intensive medical care for weeks or months, may have a different face, both in its origin and in its impact on outcome^33,34^.

Another important question is what are the factors which cause the typical hormonal changes seen in NTIS? Previous and going research in this field focuses on pro-inflammatory cytokines. From time to time various cytokines have been implicated e.g. interleukin 6 (IL-6), interleukin 1β (IL-1β), soluble interleukin-2 receptor (sIL-2R) and tumor necrosis factor (TNF)α are elevated in septic NTIS patients preceding the decrease in serum T3 and T4^35,36^. More recently some other cytokines have been found e.g. nuclear factor κB (NF-κB) causing LPS-induced deiodinase 2 increase in cell culture^37,38^. A puzzling feature of NTIS is the absence of increased serum TSH in spite of markedly decreased serum T3 and T4, pointing to altered set-point regulation of the hypothalamus-pituitary-thyroid (HPT) axis. Many possible mechanisms have been reported causing this downregulation e.g. reduced hypothalamic TRH expression and suppression of TRH in the hypothalamic paraventricular nucleus probably mediated by increased local T3 production, resulting from increased DIO2 expression in combination with decreased DIO3 expression in neurons^39,40^. Treat or not to treat NTIS has been a moot point. Current evidence suggests that NTIS may be a combination of physiologic adaptation and pathologic alteration, with no unifying benefit of treatment with thyroid hormone^41^.

In the present study volunteers belonged to all provinces and areas of Pakistan. We will recommend to use these reference values not only in Pakistan but also in surrounding countries and in overseas populations living in other countries.

The instrument we used for analysis of these hormones is the state-of-the-art random access fully automated system (Maglumi X8). CLIA is the ultimate development of time-honored ELISA plate readers, that required immense time and manpower resources with many potential sources of errors^42^.

### Conclusion

We have reported reference values of four hormones for use in clinical practice for diagnosing and monitoring of non-thyroidal illness, a very common condition present in critically ill patient.

### Recommendation

Facilities for estimation of all these four hormones on a state-of-the-art fully automated ABEI-based chemiluminescence system be made available in hospital laboratories, so that diagnosis and monitoring of NTIS is carried out effectively by the physicians.

## Data Availability

All relevant data are within the manuscript and its Supporting Information files.

## Acknowledgement

We are greatly indebted to SNIBE (Shenzhen-China) for providing reagents and consumables for this project and Pakistan Society of Chemical Pathologists (PSCP) for funding this project.

## Ethics Statement

▪ Present article reports portion of the data of a project carried out to facilitate the users of fully automated ABEI-based Chemiluminescence System Maglumi X8.
▪ By no means we report any comparison of performance of this system with any other system.
▪ No conflict of interest is declared by the authors, since no comparison of instruments was involved.
▪ This study was reviewed and approved by the Institutional Review Board of Mohi Uddin Islamic Medical College Mirpur AJ&K, Pakistan.
▪ Written informed consent of the volunteers was obtained for the study
▪ To protect privacy of the volunteers all laboratory results were blinded by allotting unique numbers to the blood specimens. The lab results of outliers, however, were unblinded for repeating and referral to the related physicians for management.

